# A roadmap for navigating partner engagement in community-based autopsy studies: Lessons from the field in rural KwaZulu-Natal, South Africa

**DOI:** 10.1101/2024.11.14.24317019

**Authors:** Alison Castle, Gugu Shazi, Threnesan Naidoo, Ashendree Govender, Nceba Gqaleni, Emily B. Wong, Collins Iwuji, Kobus Herbst, Adrie JC Steyn, Mark J. Siedner

## Abstract

**Background:** The measurement of cause-specific mortality is critical for health system planning but remains a challenge in many low-resource settings due to societal, legal, and logistical barriers. We present a co-development process with community members for the design and implementation of an autopsy program to improve cause of death data in a historically underserved population.

**Methods:** We sought to develop an autopsy program at the Africa Health Research Institute (AHRI) Health and Demographic Surveillance Site (HDSS). The project proposes to obtain consent from families of deceased adults, to perform diagnostic autopsies by a trained pathologist, and to process samples to determine causes of death. Prior to launching the program, we engaged key partners in learning their perspectives about such a program and understanding the landscape of challenges needed for successful implementation. Herein, we describe lessons from interactions with these partners, including 1) the AHRI community advisory board (CAB), 2) the South Africa Department of Health (SA DoH), 3) local traditional authorities, 4) funeral home personnel, 5) traditional healers, and 6) healthcare providers. We also detail the development of community outreach efforts used to inform the public about the program.

**Results:** The partners provided valuable feedback on the study design and informed us of issues that needed to be addressed: community concerns about organ retention and sale (CAB), implications of how autopsy findings could spur litigation and erode trust in healthcare providers who determined alternate causes of death (SA DoH), a cultural practice that conflicts with the autopsy procedure (traditional healers), the need to educate families before they engage with funeral businesses (funeral homes), and enhancing our death referral network through healthcare providers. This led to protocol changes and an adapted community engagement strategy, which included educating healthcare providers, hosting community dialogs, broadcasting radio advertisements, and developing a film to describe autopsy procedures to families considering participation.

**Conclusions:** We present a comprehensive model of partner engagement for a community-based autopsy program in South Africa, leading to the co-development of a program that incorporates local customs around death while promoting buy-in and support from the government, civil society, and medical partners.

## Introduction

Accurate identification of the leading causes of death is crucial for effective health system planning and resource allocation. In rural South Africa, as in other low- and middle-income countries, the collection of accurate mortality data is limited by a scarcity of postmortem data.(1) Modeling studies such as the Global Burden of Disease Study, which use risk factors to estimate mortality in areas with limited data, suggest that HIV/AIDS is a leading cause of death in the country.(2) Yet, more recent data suggest that this trend may be changing.(3) The widespread adoption of antiretroviral therapy has led to a notable decline in deaths due to HIV.(3) However, the leading causes of death emerging from this epidemiological shift remain obscure, largely due to incomplete or inaccurate death registries that do not reflect directly measured causes of mortality.(4, 5)

In settings where cause-of-death information is limited and where most deaths occur outside healthcare facilities, verbal autopsy has emerged as a common method for determining population-based, cause-specific mortality.(6) Verbal autopsies are used by multiple demographic health and surveillance sites and have been shown to have favorable validity for accidents, trauma, and infectious disease causes of death.(7) However, for deaths related to cardiovascular disease or cancer, verbal reports have poor sensitivity and specificity, particularly in settings where diagnostic infrastructure for those conditions is lacking.(7)

To address this knowledge gap, we are developing a pathologic autopsy program within a rural Health and Demographic Surveillance Site (HDSS) in KwaZulu-Natal, South Africa, encompassing 150,000 individuals. The objectives of the pathologic autopsy program are 1) to determine the leading causes of death among adults in rural KwaZulu-Natal in the era of widespread antiretroviral therapy availability and declining HIV mortality(8, 9); 2) to evaluate the validity and determine the optimal use of verbal autopsy-based tools; and 3) to provide the health department with high-quality cause of death and burden of disease data to inform policy. Although complete diagnostic autopsies are considered the gold standard for determining the cause of death, such practices have been historically challenged in resource-limited settings due to the lack of histologic laboratory infrastructure and skilled personnel.(6) Our initiative in South Africa aims to overcome these challenges by leveraging a robust research infrastructure, including diagnostic testing, tissue storage, and the expertise of trained pathologists and prosectors, in partnership with a longstanding HDSS survey with comprehensive risk factor data.(10, 11)

In preparation for the introduction of this program, we met with a diverse set of public, civil society, and medical stakeholders to learn about preferences for and challenges to its implementation. We recognized the many cultural and religious beliefs that often pose challenges to conducting postmortem examinations, which involve complex discussions with family members after the loss of a loved one and examination of the external and internal body with invasive sampling to obtain tissue for microscopy.(6) In South Africa, many sociocultural barriers to autopsy, such as gendered power dynamics, traditional and religious belief systems, and a lack of trust in the healthcare system, have been reported.(12) Herein, we describe feedback provided by program partners that we hope will offer insights into the establishment of autopsy programs in similar contexts.

## Methods Study

### Setting

This work was conducted at the Africa Health Research Institute’s (AHRI) Health and Demographic Surveillance Site embedded within the Hlabisa subdistrict of KwaZulu-Natal, South Africa. Since the initiation of AHRI’s verbal autopsy program, we have observed over 2,550,000 person-years and recorded over 24,000 deaths.(13) Annual household surveys capture data on migration, household wealth, use of healthcare services, and clinical testing for prevalent diseases such as HIV, tuberculosis, hypertension, and diabetes. The population remains one of the world’s most intensely HIV-affected regions, with more than 34% of the population living with HIV.(10) Due to tremendous public health efforts that have increased access to antiretroviral therapy, more than three-quarters of those with HIV have controlled disease with suppressed viral loads.(9) Most residents live beneath the South African poverty line in rural or peri-urban areas, with an overall life expectancy of 56.1 years.(14) Within this community, medical pluralism is common, and many individuals consult traditional healers before seeking care at Department of Health facilities.(15, 16)

### Initial Proposed Autopsy Study Design by the Research Team

The outline of a community-based pathologic autopsy program was presented to members of the community and was iteratively refined (**Figure 1**). Eligible participants for the autopsy study include deceased residents of the HDSS aged 18 years or older. The pathologic autopsy program would be offered to families of all deceased adults who meet eligibility criteria. Recruitment would be facilitated through funeral homes. Upon notification of a death from the funeral home or interested family, the project coordinator would contact the family and establish an in-person meeting to discuss the study. Informed consent would be obtained from the next of kin in accordance with the South African National Health Act (17) at a location chosen by the family, which could be the funeral home or the family’s residence.

**Figure 1:**
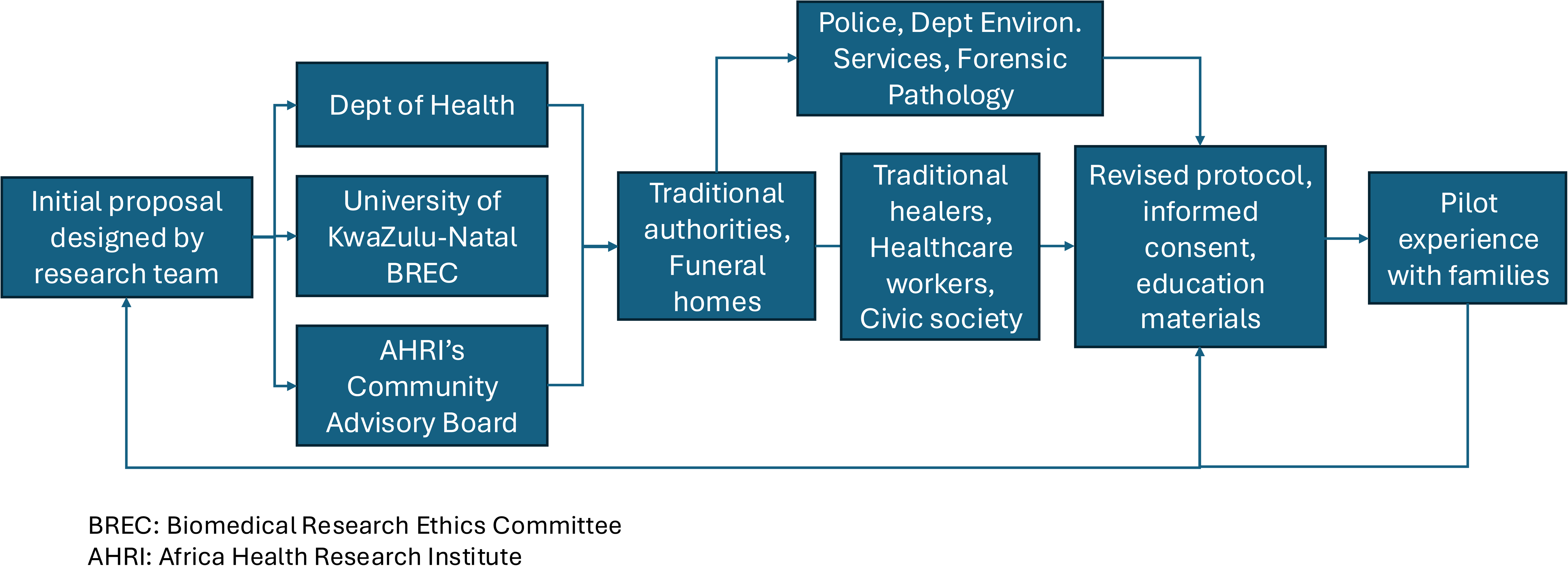
Collaborative Framework for Protocol Development and Implementation. This diagram outlines the comprehensive approach taken by the research team at the Africa Health Research Institute (AHRI). It highlights the initial proposal design with the Department of Health, University KwaZulu-Natal Biomedical Research Ethics Committee, and the community advisory board. It then shows progression through the extensive community and institutional engagement process involving traditional authorities, funeral homes, healthcare workers, civic society, the police department, environmental services, and forensic pathology. The figure illustrates the iterative process of refining protocols, informed consent, and educational materials through pilot experiences with families and community partners.

### Additional proposed study procedures

Following receipt of informed consent from the family, the study pathologist would perform the autopsy at the affiliated funeral home, adhering to any family requests regarding the extent of the procedure or specific organ sampling for analysis. Tissue processing and storage would be performed at AHRI’s laboratory in Durban. Families would receive autopsy results after 12 to 16 weeks, which include the most likely cause of death and contributing causes. A verbal autopsy, which occurs at 12 weeks after a death within the HDSS protocol, would then be conducted at a separate time with the closest relative before disclosure of the final pathologic autopsy results.

### Partner Engagement Methods for Iterative Refinement of the Autopsy Study

This study was designed to incorporate the perspectives and expertise of diverse groups, including the Africa Health Research Institute’s Community Advisory Board (CAB), the University of KwaZulu-Natal’s Biomedical Research Ethics Board (BREC), the Department of Health officials, local traditional authorities, funeral home personnel, traditional healers, the South African Police, Civil Society community leaders, and healthcare providers associated with AHRI and the Department of Health.

Our engagement approach varied by partner type to maximize accessibility and participation. The AHRI Public Engagement Team facilitated meetings with key partners such as traditional authorities, the Department of Health, and civil society leaders. Our engagement with traditional healers was facilitator by the Traditional Medicine Faculty at AHRI. For funeral home engagement, we conducted a comprehensive search to identify every funeral home within the region, followed by in-person appointments with each facility.

Recommendations for key concepts to address and findings from each partner, as well as resulting protocol adaptations made in response to their input, are summarized below and in **Table 1**.

**Table 1:** Partner Engagement and Protocol Adaptations in Community-Based Pathologic Autopsy Program.

| Key Entity/Partner | Topic Areas Covered | Key Lessons Learned | Implications and Protocol Adaptation |
| --- | --- | --- | --- |
| Community Advisory Board (CAB) | Burial practices<br>Beliefs about autopsy<br>Cultural appropriate messaging | Burial within ~7 days of death<br>Witchcraft beliefs<br>Misconception is that forensic autopsies remove organs for traditional medicine, replaced with cotton | Ensured cultural sensitivity in outreach strategies, clarified medical objectives of autopsies, refined communication strategies to address misconceptions forthright |
| Department of Health | Distinguishing between forensic and natural death policies<br>Trust of healthcare providers<br>Medical-legal implications | Need to align with public health goals and regulations | Confirm only natural deaths were eligible, delineated protocols to not overlap with unnatural death referrals, clarified study scope to not implicate medical malpractice |
| Local Police Force | Referral pathways for unnatural deaths | Immediate notification to police if sudden death or suspect foul play | Established protocol for transferring cases to police if unnatural death is suspected |
| Local Traditional Authorities | Acceptance of program | Required endorsement for any study in the region | Gained approval to continue with autopsy study |
| Funeral Home Association | Engagement and partnership<br>Use of facilities<br>Facility compliance<br>Referral strategies | Concerns about the role of funeral home staff communicating about autopsies, legal implications of conducting autopsies within facilities | Shifted to direct community outreach and community referrals instead of relying on funeral home staff to convey study, formalized collaboration through contracts, rented morgue space |
| Traditional Healers | Acceptance of program<br>Traditional practices | Significance of traditional ritual ukushashazela | Adapted communication and consent processes to include provisions for traditional practices |
| Healthcare Providers' Partnership | Education of procedure benefits<br>Referral of appropriate families | Involving healthcare providers is key for broadening reach | Referral pathways primarily through healthcare professionals |
| Civil Society Meeting | Acceptance of program | Required endorsement for any study in the region | Gained approval to continue with autopsy study |

### Africa Health Research Institute’s Community Advisory Board (CAB)

We initiated our engagement process by conducting meetings with the Africa Health Research Institute’s Community Advisory Board (CAB), an official oversight body that evaluates study designs in parallel to the Biomedical Research Ethics Committee (BREC). The CAB, which convenes monthly, plays a pivotal role in representing community interests and ensuring ethical research practices. The 25 members are elected every three years and receive comprehensive three-day training on roles, responsibilities, and AHRI’s research activities. The CAB enables community members to express any questions or concerns about the institute’s research prior to and throughout the duration of community-based projects.

The CAB was instrumental in educating our research team about various cultural, traditional, and community norms that could influence engagement with the autopsy program. They brought to our attention considerations about local beliefs, such as ukuthakatha or witchcraft, and how they might intersect with our research objectives. For example, if a person was believed to have been the target of witchcraft, family members may seek information about how it may have contributed to the participant’s demise. We addressed this concern by including language in our consent forms and study information materials that the procedure would only detect the medical diagnoses that may have contributed to death.

The CAB was supportive of the study’s objectives. However, they emphasized the need to time autopsy procedures considering local burial customs, which typically occur within a week of death. Acknowledging this, we ensured that our consent process explained that we would strive to accommodate the time-sensitive burial proceedings and that families may want to decline if there was a risk that it would delay burial plans. We also clarified that there would be no additional costs to the family if they chose to wait until the autopsy procedure could be completed before burial. Additionally, the CAB members raised a common misconception that forensic pathologists remove all organs during autopsies and replace them with cotton, which conflicts with the local tradition of burying a person “whole”.(12) We proactively addressed this by clarifying that we do not replace organs with cotton or remove full organs in our community outreach methods. Finally, the CAB helped us refine communication strategies, such as the wording used in radio broadcasts and other public communications, to ensure a high degree of cultural sensitivity (**Figures 2,3)**.

**Figure 2:**
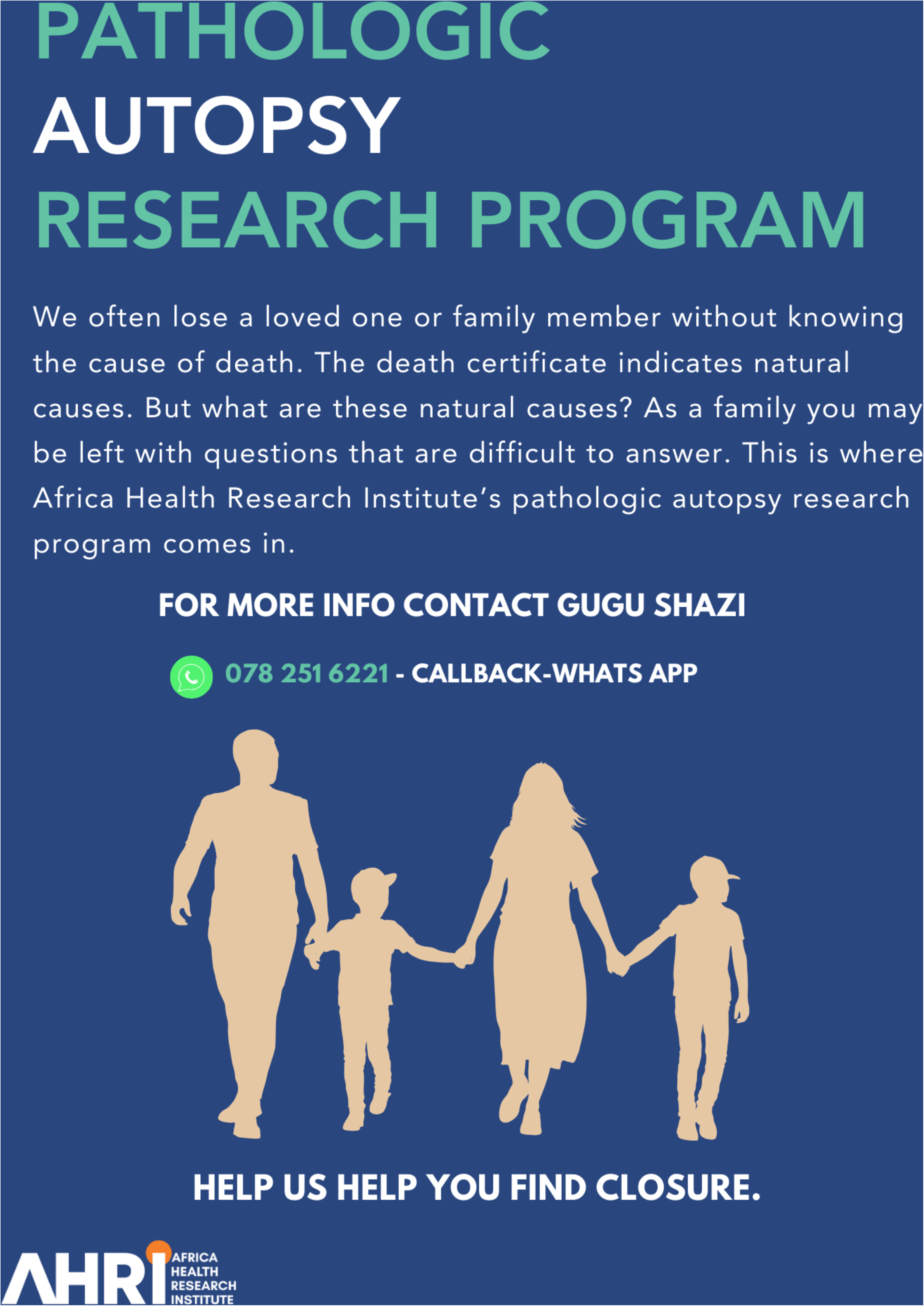
Participant-facing flyer for Study Outreach and Engagement. (English version) This flyer, designed by the Africa Health Research Institute’s Pathologic Autopsy Research Program, is addressed to families who may be left with questions after a family member is deemed to have died from natural causes. It raises awareness about the autopsy program goals and invites community members to seek more information through the contact details for the program coordinator.

**Figure 3:**
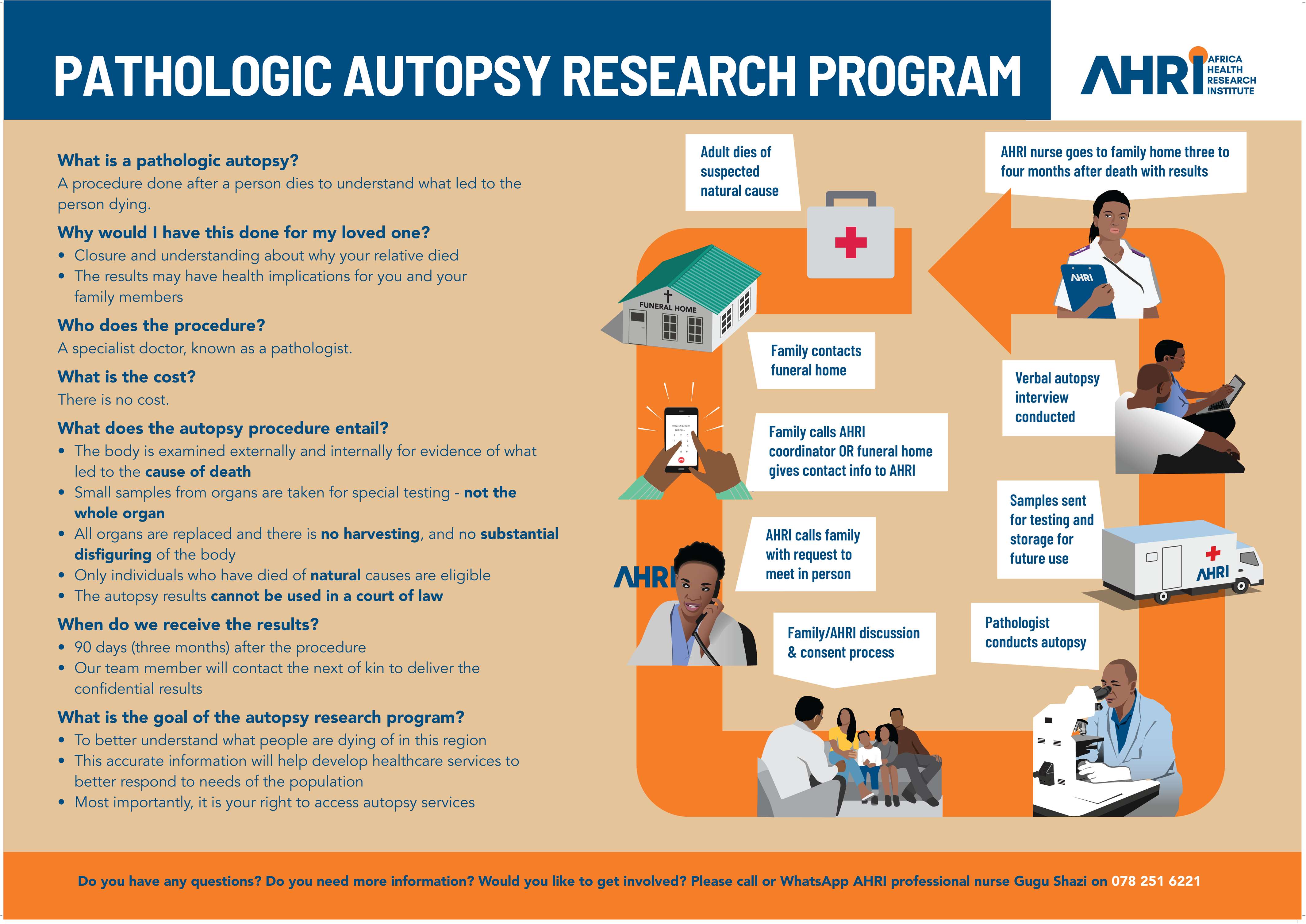
Educational Flyer for the Pathologic Autopsy Program (English version) This poster explains the concept, purpose, and process of a pathologic autopsy. It addresses common questions raised by our community partners. A diagram illustrates the step-by-step procedure and concludes with the contact information of the study coordinator.

### University of KwaZulu-Natal’s Biomedical Research Ethics Board (BREC)

The Biomedical Research Ethics Board provided ethical review of our protocol and consent forms. Feedback from this group resulted in our study adopting a separate consent form for tissue storage and future use of biological samples. Additionally, to ensure compliance with confidentiality standards, the input from the ethics committee led us to establish an independent consent process for collecting contact information from potentially eligible and interested families at the funeral homes. To do so, we created a document capturing the name and contact information of an interested family member, which was provisioned in each of the funeral home businesses. We were also asked to provide our study personnel with specialized training in 1) ethical consent practices for the recruitment of recently bereaved families and 2) the management of emotional stress among study staff. The training was provided by the staff psychologist at AHRI psychologist. Standing team debriefing sessions with the verbal autopsy nurses and pathologic autopsy nurse were scheduled.

### Department of Health

Discussions with the Department of Health were crucial to align the study procedures with local health regulations. During these meetings, district and provincial officials were consulted to delineate the scope of our autopsies, ensuring that they were clearly defined as examinations for natural causes of death and thus would not interfere with forensic pathology services, which are legally mandated for deaths suspected to be unnatural.(18) Natural deaths are defined as deaths related to internal bodily events not influenced by external occurrences, such as suicide, homicide, or trauma. Furthermore, if the study pathologist identified findings concerning an unnatural death, the deceased would be immediately transferred to police custody as sanctioned by the Department of Health. This change was adapted to our protocol as well as to AHRI’s Memorandum of Agreement with the Department of Health.

Several additional unforeseen concerns also surfaced in discussions with the Department of Health. For example, they queried whether autopsy results could be used in legal action against healthcare providers for misdiagnosis or medical negligence, particularly if a patient was treated for one disease, such as tuberculosis, but was later found through autopsy to have died from a different disease process. We addressed this issue by clarifying the scope of the study, emphasizing that the primary aim is to advance medical understanding and not to provide evidence for malpractice litigation. We also revised our communication and consent documents to state that the clinical research findings should not be used in a court of law to ensure that participant families were not engaging in this research program for purposes of litigation. We reinforced the role of autopsies in improving diagnostic accuracy and healthcare quality, underlining that the study would ultimately strengthen the community’s health system by providing insights into disease patterns.

### Community Police Forum

Discussions with the police force informed our team of the appropriate referral pathways for unnatural death reporting. Although participants are only eligible for our study if they are deemed to have died from natural causes, there may be potential cases where the study pathologist identifies findings suspicious for an unnatural cause of death. We learned the necessary steps needed to transfer the participant to police jurisdiction and amended our protocols to account for this possibility.

### Local Traditional Authorities

In KwaZulu-Natal, the endorsement and support of local traditional chiefs are prerequisites for community-based research, but particularly for studies addressing sensitive topics, such as postmortem examination. We approached local chiefs to obtain their input and advice on our study design. In our meetings, we described the study’s objectives, potential benefits to the community, and the approach we would adhere to regarding cultural practices surrounding death. Through open dialog and answering their questions, we received their approval to proceed.

### Funeral Home Collaboration

We conducted a series of face-to-face conversations with funeral home managers and staff, which revealed insights that influenced our outreach and operational strategies. Initially, these dialogs aimed to explore the potential for partnership and gauge the level of interest in the study. However, as discussions progressed, funeral home managers expressed concern about being the primary point of contact for families to learn about the autopsy study. They stressed the need for the research team to proactively educate the community to alleviate the burden on funeral home staff of conveying sensitive information about autopsy practices. This feedback led to a strategic shift in our approach, prioritizing direct community outreach to introduce the study and its objectives before family engagement with funeral services.

Furthermore, funeral home managers were apprehensive about the legal implications of conducting autopsies on their premises. To address these concerns, we sought approval to perform autopsies within legally licensed funeral homes from representatives of the Department of Health Forensic Pathology Division and the Department of Environmental Services. Additionally, we learned that among the 17 funeral home businesses in the region, the majority shared resources, with three common morgues used collectively between them. To formalize the collaboration, we developed a contract with each funeral home, detailing the relationship between AHRI and the funeral home entities. No financial incentives were offered to the funeral homes to ensure ethical integrity and prevent the coercion of program participation. Instead, we opted to rent morgue space at the standard rate, a cost equivalent to what smaller funeral businesses would incur when utilizing the morgues of larger establishments.

Through these engagements with funeral home staff, it became evident that, whereas there was an interest in medical causes of death, they felt most families seeking autopsies did so under the suspicion of unnatural causes, such as poisoning, witchcraft, or foul play. This distinction was important in tailoring our communication and consent processes to ensure that families were in line with our study goals to determine the natural causes of death in the community.

### Traditional Healers

Recognizing the influential role of traditional medicine in the community’s healthcare decisions, we engaged with traditional healers to seek their impressions and insights into an autopsy program. The traditional healers taught us more about the local community practice of ukushashazela, in which families perform a ritual involving the slaughter of a goat to “close the wounds” for a relative buried without all organs or tissues.(19) The traditional healers advised that autopsies could be culturally accepted if provisions for ukushashazela were available to families who practice it. Considering this, we refined our autopsy communication and consent processes to mention such cultural practices. After addressing their concerns and providing detailed explanations about the study, we sought support from traditional healers in recommending autopsies to appropriate families, with confidence that our procedures would be respectful of local customs.

### Healthcare Providers

We also conducted a series of group discussions and educational sessions with nurses, community healthcare workers, peer navigators, and physicians affiliated with AHRI and the Department of Health. This included engagement with 231 healthcare providers affiliated with 11 primary health care clinics and the district referral hospital. These sessions served to inform healthcare professionals about the study procedures, answer any queries, and encourage them to refer interested families who met the criteria to our study team.

### Civil Society Leaders

Within the uMkhanyakude district, civil society meetings include diverse representations of community leaders. These include members from the Department of Health, social workers, Non-Governmental Organization (NGO) representatives, and District and Local AIDS Council members. These meetings served to further disseminate the objectives of the pathologic autopsy study and receive approval for study implementation.

### Community Engagement and Outreach

The above partner meetings helped to inform a comprehensive community outreach and sensitization plan. Goals for the program included informing the public about 1) the value of cause of death determination both individually and for the community; 2) the steps, nature, and timing of autopsy procedures; and 3) contact information for interested parties. We disseminated the program in the following ways:

### Entertainment-Education Road Shows

The study team participated in the HDSS monthly road shows that combine education with entertainment, a method that has proven effective in previous AHRI studies. These events allowed direct interaction between the research team and members of the community, facilitating a dynamic exchange of information. The road shows were designed to be engaging and informative, covering specifics of the autopsy study procedures, including data collection processes and how results would be disclosed to participant families.

### Radio Broadcasts

To extend our reach across the HDSS community, we partnered with two local radio stations, broadcasting a 30-second informational clip inviting listeners to tune into an hour-long program. These sessions, led by the study’s project coordinator in isiZulu, were scheduled regularly and allowed us to address community questions in real time. Many of the questions reflected community beliefs and concerns already raised during the CAB meetings.

### Community dialogs

The Public Engagement Unit at AHRI conducts community dialogs in isiZulu to address myths and concerns regarding research activities within the HDSS. The dialogs are scheduled to align with the HDSS’s surveys, targeting areas where surveys are to be conducted the following week. Although all community members from these areas are invited to participate, typically only 10-20 members attend the discussions, along with the project coordinator from the Pathologic Autopsy Program. These gatherings take place in accessible outdoor spaces or homesteads. The number of dialogs held is contingent upon the size of the area and the nature of the concerns raised, ranging from two to three sessions.

### Verbal Autopsy Infrastructure

We leveraged the existing verbal autopsy infrastructure for reporting demographic events. For example, any individual may call AHRI to report births, deaths, migrations, etc., within the HDSS. Individuals who report these events to the AHRI call center are offered airtime reimbursements, ensuring that the cost does not hinder communication. We integrated the pathologic autopsy program into this established system to enable more rapid notification by our project coordinator. Our coordinator would then reach out to the notifying individual and contact the relatives with a recent death to inform them about the autopsy study and assess eligibility. This enabled more targeted information sharing to families who may be interested in the autopsy program.

### Dissemination of Autopsy Program Educational Adverts

Our outreach materials included flyers and posters (**Figure 2**, **Figure 3**). Flyers are distributed to the community at the Department of Health clinics, community events, and funeral homes. Posters, which more comprehensively detail the autopsy process, were strategically placed within funeral homes to educate and inform visitors.

### Video-Based Consent Process

We planned to obtain consent through in-depth face-to-face discussions, ensuring that families were well informed before participation. We developed a consent video presented in isiZulu, featuring the project coordinator, the Community Advisory Board Chair, the study pathologist, and the study prosector. This five-minute video (**Supplemental 1**) proved to be a crucial tool for clearly explaining the study’s goals, the pathologic autopsy and verbal autopsy components, and the tissue storage process in our pilot trial. The film’s effectiveness in conveying complex information succinctly made it an invaluable asset not only during the consent process but also during partner meetings.

### Feedback from individuals with recently deceased family members who declined participation

As community knowledge about the autopsy program increased, several families were referred to discuss the procedure. Initially, the first 10 families declined to consent for autopsy. Reasons for declined enrollment included the following: 1) a death certificate had already been issued, and the family was content with the diagnosis; 2) the timing of the autopsy procedure, which was conducted weekly on Mondays or Tuesdays by our study pathologist, did not align with the funeral ceremony preparations; 4) family members were not the next of kin, or they requested permission and consensus from others; and 5) the return of results 12-16 weeks later was deemed untimely. To address these barriers, we modified our study approach. For example, we partnered with a private physician in the community responsible for completing death certificates to add preliminary data found on autopsy examination to the death forms. We have expanded the number of days the study pathologist is available to conduct autopsies to facilitate family preferences for funeral proceedings and funeral home obligations. Finally, in our protocol, we sought written consent from the designated next of kin but strongly encouraged the consensus of the entire family to maximize joint decision making.

## Discussion

This work highlights the importance of community engagement and partner feedback in the establishment of a community-based pathologic autopsy program within rural South Africa. Proactive and culturally sensitive strategies, codeveloped with community members and civic leaders, are requisite in areas where autopsies are not commonly conducted due to sociocultural, religious, and logistical barriers. Our meetings with partners provided a number of key insights, such as the need for referral pathways that are independent of funeral homes, educational materials that address concerns among community members, the need to integrate a multistep process to confirm family motivations for pursuing autopsy without suspected foul play, the need to ensure the alignment of autopsies with traditional practices (in our case with that of ukushashazela), and the ability to provide flexibility for when the procedure is conducted to align the procedure timing with planned funeral proceedings.

This work was motivated by reports from the region about hesitancy surrounding pathologic autopsy procedures. For example, research in Tanzania found that community attitudes toward autopsies were influenced by beliefs about witchcraft and concerns regarding the disturbance of the body after death, akin to feedback raised during our partner interviews.(20) In Zambia, one study demonstrated that families often decline neonatal autopsies, with 43% citing them as futile since the diagnosis should have occurred prior to death, rendering postmortem findings of no benefit.(21) Additionally, more than one-quarter of those who refused did so because a death certificate had already been issued and arrangements to transport the body had been made and could not be delayed.(21) In contrast, a study conducted across Gabon, Kenya, Mali, Mozambique, and Pakistan found that interviews with family members in the first 24 hours after death were easier to conduct than those at later stages.(22) This group also found that the acceptability of an autopsy program required early community education and staff preparedness to be able to appropriately address emotional trauma and grief while ensuring a comprehensive consent process. Based on this information, we sought to meet early and often with relevant community members to understand and address concerns they may have and seek solutions to garner support.

Our experience provides a roadmap for researchers and healthcare practitioners aiming to implement similar community-based pathologic autopsy programs in other settings. We took away three key lessons from our program development through engagement with a diverse set of community members, including the Department of Health, local traditional leaders, community advisory boards, the police force, civic society members, healthcare workers, and funeral home personnel. First, engaging with community members to understand cultural beliefs and misconceptions about autopsy is an instrumental first step. While other studies have explored perspectives from the relatives of recently deceased persons, (22–26) we started by holding group discussions with community members not currently experiencing bereavement to gain an understanding of traditional and cultural practices, appropriate messaging techniques, and delve into the nuances of burial practices. Feedback indicated that the timing of the autopsy procedures was critical. Aligning the study to accommodate local burial customs and family preferences for funeral proceedings led to expanding the availability of the study pathologist and adjusting the consent process to encourage family consensus. Furthermore, as autopsy programs are not successful without input and buy-in from local traditional and civic society groups, scientific modifications may be required after their perspective is gained. Our experience underscores this point; adaptation of our communication and consent processes was necessary after gaining a deeper understanding of ukushashazela. Incorporating this cultural aspect in our approach helps ensure that the program is respectful of and responsive to the values and beliefs of the community we aimed to serve.

Second, because the legal and regulatory frameworks governing postmortem examinations vary significantly by country and jurisdiction,(27) meeting with legal experts and local health authorities is essential to ensure compliance with autopsy procedures. We were first aware of these legal requirements through funeral home engagements. Autopsy studies using funeral home facilities have previously been met with challenges.(28) The private business owners raised concerns about legal implications, which ultimately led us to formalize business contracts, rent morgue space at standard rates, and engage with the Department of Environmental Services to oversee facility compliance to ensure that the funeral home facilities met standards for autopsy. We also engaged the Forensic Pathology Division of the Department of Health and police force to ensure that our protocols aligned with the policies for unnatural death referrals.

Finally, community sensitization is a prerequisite step ahead of community-based autopsy programs. Promoting community buy-in requires transparent two-way communication about the program’s goals, processes, and potential benefits. We employed a multifaceted outreach strategy with the goal of broadly reaching all members of the target population, including entertainment-education road shows, radio broadcasts, community dialogs, and the use of video-based consent processes. The use of video in the informed consent process for medical research, including autopsy studies, offers notable benefits compared to traditional text-based approaches. Video-based consent has been shown to lead to enhanced enrollment and greater diversity among study participants.(29) This finding suggests that video consent can assist in making complex medical information more accessible and understandable. For our autopsy consent process, we provide a clear explanation of the procedures through video aids to dispel misconceptions that could impede consent. The video details the procedure comprehensively (albeit without images of a deceased person) and features key individuals such as the pathologist and project coordinator to humanize and clarify the process for potential family members. Additionally, by following an example family through the stages of the autopsy process, the videos attempt to offer a relatable and transparent narrative that can be particularly beneficial when overcoming language barriers or emotionally sensitive subject matter.

### Conclusions

The establishment of a community-based pathologic autopsy program requires a comprehensive community-engaged approach. By following this structured roadmap, researchers can navigate the complexities of implementing similar programs, thereby enhancing mortality data and informing public health strategies in underserved populations. Our experience illustrates that with careful planning, respectful engagement, and adaptability, it is possible to overcome barriers to implementing an autopsy program and contribute valuable insights into the causes of death in communities where such data are scarce.

## Data Availability

Data sharing is not applicable to this article, as no datasets nor interview guides were generated or analyzed during the current study.

## Declarations

### Ethics Approval and Consent to Participate

This study received ethical approval from the Biomedical Research Ethics Committee (BREC) at the University of KwaZulu-Natal (reference: BE290/16). The committee endorsed the publication of our manuscript that reports feedback from stakeholder engagements as it aligns with the approved protocols and enhances the social value of our work, with no further action required from the Principal Investigator to obtain written informed consent before publication. Therefore, written informed consent was not obtained from all stakeholder participants. This approach was approved by the BREC under specific conditions that ensure compliance with ethical standards.

### Consent for Publication

Written consent for publication of Supplemental 1 Consent Video was obtained from all participants.

### Authors’ contributions

GS, AC, TN, and NG conducted the partner engagement and community outreach activities. GS, AC, TN, AG, EW, CI, KH, AS, and MS contributed to the design of the study. AS obtained funding for the program. All the authors have read and approved the final manuscript.

## Acknowledgments

The authors would like to thank all the community partners and families who provided their feedback on the development of this study.

